# Cohort Profile: Generation Victoria (GenV)

**DOI:** 10.1101/2025.08.21.25333171

**Authors:** Elizabeth K. Hughes, William Siero, Alisha Gülenç, Anna Fedyukova, Susan A. Clifford, Tony Frugier, Jatender Mohal, Naomi Schwarz, Daisy A. Shepherd, Melinda Barker, Zeffie Poulakis, Joanne M. Said, Natasha Zaritski, Sharon Goldfeld, Richard Saffery, Melissa Wake

## Abstract

**Key Features:**

- Generation Victoria (GenV) is a whole-of-state research platform established to improve child and adult health, development and wellbeing through discovery and interventional research. It comprises participant-provided and service-collected data and biosamples, with planned 5-6 yearly child and parent phenotypic waves.
- All children living in Victoria, Australia, born between 4 October 2021 and 3 October 2023 and their parents/guardians are eligible for GenV’s ‘*Cohort 2020s’*. Recruitment remains open.
- We describe the GenV Cohort ‘Foundation Set’ recruited to 1 May 2025. Of eligible children, 29.7% had joined *Cohort 2020s* (43 945 children; 64 106 parents/guardians). Together with an ‘*Advance Cohort*’ (born December 2020-October 2021), we recruited 49 783 children and 74 270 parents/guardians (48 213 birth parent/mothers; 25 428 fathers; 314 other parents/guardians); 761 (0.6%) had withdrawn.
- *Cohort 2020s* birth mothers/parents were aged 15-54 years, 23% lived in regional/rural areas, and 25% spoke non-English languages.
- Data and biosample collection includes baseline surveys, biosamples (e.g., saliva 81%, stool 16%, breastmilk 16%); optional inter-wave surveys (up to 33%); residual antenatal (up to 28%) and birth biosamples (up to 97%), and extensive linkage to services, administrative and geospatial data (up to 98%).
- Numerous collaborative research projects are under way; end-user applications will commence 2026.

## Why was the cohort set up?

Generation Victoria (GenV) was created to help solve pressing challenging facing today’s children and adults – rising rates of chronic disease, falling life expectancy, emergent physical, mental, and educational crises, and widening inequity [1–7]. Despite decades of research investment, non-communicable diseases (often rooted early in life) continue to rise. This reflects a persistent gap between discovery and real-world population impact.

To close this gap, prevention research needs very large, inclusive, long-term cohorts that can test how new approaches work in the real world. Lack of such contemporary mega-cohorts curtails the ability to translate data into population-level wisdom and action.

GenV was designed meet this need as a single, statewide, open science platform that integrates discovery and interventional research within an entire population [8]. It combines features of a birth cohort and a panel study, creating large, parallel cohorts of children and parents. These cohorts will be continuously refreshed to reflect Victoria’s changing population, policy and environment. By linking existing and GenV-collected data (including biosamples and phenotypes) and enabling embedded large trials, GenV acts as a real-world testbed for scalable solutions to improve prediction, prevention, equity, affordability and outcomes.

Led from the Murdoch Children’s Research Institute (MCRI), with support from The Royal Children’s Hospital Melbourne and the University of Melbourne, GenV is developing as an international research asset funded to date by the Victorian State Government, competitive grants, and philanthropy (see Funding statement).

## Who is in the cohort?

GenV is a statewide population-based cohort based in Victoria, Australia, designed for both discovery and intervention. The statewide *Cohort 2020s* is open to all children residing in Victoria with a birth date 4 October 2021 to 3 October 2023, and their parents/guardians. Recruitment to Cohort 2020s is open indefinitely, including to those moving into Victoria. A smaller *Advance Cohort* comprising a non-representative sample of children born 5 December 2020 to 3 October 2021 and their parents/guardians, enabled fine-tuning and scaling of logistics ahead of statewide rollout. Recruitment to the Advance Cohort ended in June 2022.

This profile describes both cohorts as recruited during the newborn and post-newborn periods to 1 May 2025 (the *Foundation Set*). While this phase resembles a traditional birth cohort, GenV’s ongoing design also has features of a panel: whole-population representation, continuous recruitment to reflect migration and demographics, and ongoing relevance across two generations.

Throughout the eligible birth period, GenV ascertained births daily via the statewide birth records generated by the Victorian Infant Hearing Screening Program (VIHSP). Most families were approached face-to-face in hospital shortly after birth, with those missed in hospital contacted to join by phone or via an online self-guided form sent by email or SMS. GenV continues to encourage eligible families to join Cohort 2020s by raising awareness via children’s services, community groups, registries and collaborative studies. Full recruitment and data collection procedures are described in the published GenV protocol [8].

### Cohort Recruitment

Figure 1 depicts the recruitment flow of the GenV Cohort 2020s and combined cohorts to 1 May 20205. For this profile, ‘birth mother/parent’ was defined as a person who gave birth to the index child, regardless of gender identity or guardianship status (e.g., including non-binary birthing parents, surrogate mothers). ‘Father’ was defined as a person who identified as male and as a parent or father to the index child (including step and adoptive fathers). ‘Other parent/guardian’ included all other types such as grandparent, donor, same-sex partner of birthing mother, and intended mother of a surrogate child. Collectively, all types are referred to as parents or parents/guardians.

**Figure 1:**
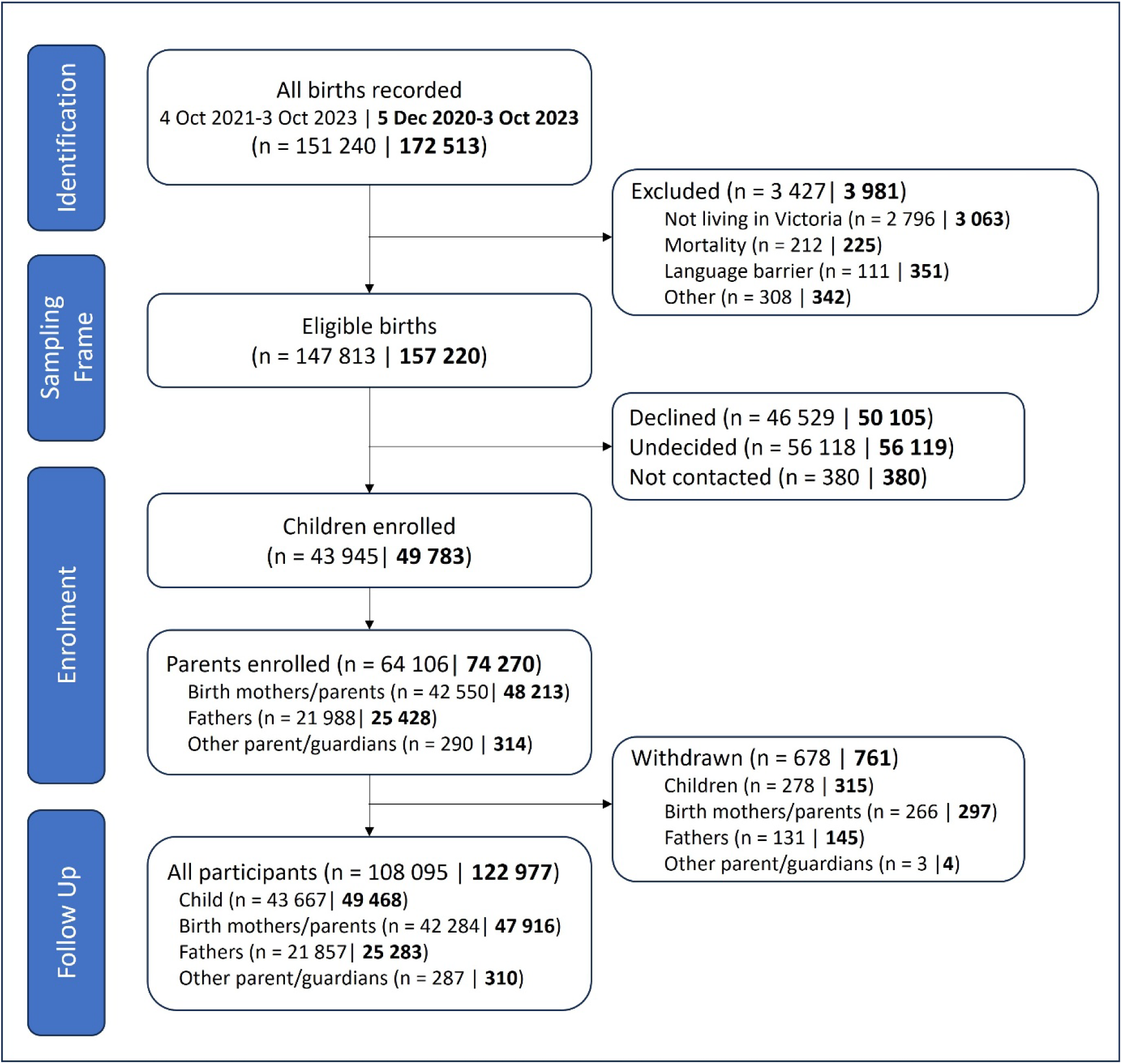
Participant recruitment flow for Cohort 2020s and combined Advance Cohort and Cohort 2020s as of May 2025 Plain text is Cohort 2020s only | Bold text is Advance Cohort and Cohort 2020s combined. Alt Text: Graphical representation of the four phases of the cohort recruitment – identification, sampling frame, enrolment, follow up.

For Cohort 2020s, 99.7% families of known eligible births had been invited in-person or by phone/email/SMS; 61.4% of these reached a decision to join or declined while the remainder did not respond/decide. In total, 108 051 participants were enrolled including 43 945 children and 64 106 parents/guardians (42 550 birth mothers/parents, 21 988 fathers and 290 other parents/guardians). Based on recruitment of at least one child and parent/guardian, this represents a response rate of 29.7% of all known eligible births, or 48.6 % of all those who reached a decision.

By 1 May 2025, 124 053 participants had enrolled in the Advance Cohort or Cohort 2020s (49 783 children, 74 270 adults), with 761 (0.6%) having withdrawn.

### Cohort Demographics

Tables 1 and 2 describe the cohort demographics for birth mothers/parents, fathers, and children, excluding those withdrawn prior to analysis. Supplementary Table 1 describes the demographics of the GenV Parent Cohort inclusive of all parent/guardian types regardless of gender or relationship (e.g., non-birthing mothers, grandparents). Supplementary Tables 2 and 3 describe cultural, linguistic and socioeconomic profiles of the Advance Cohort and Cohort 2020s in further detail.

**Table 1.**
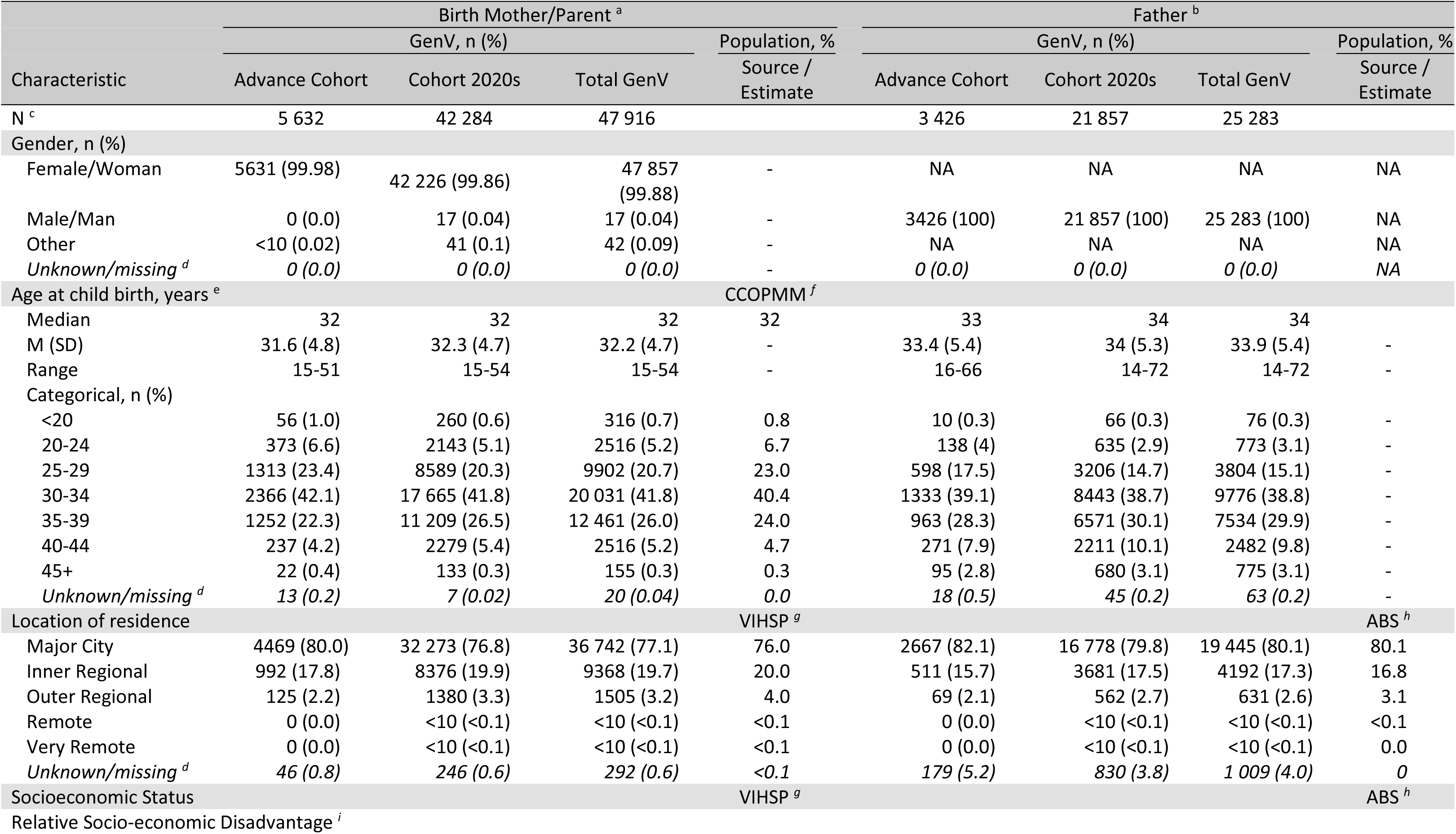
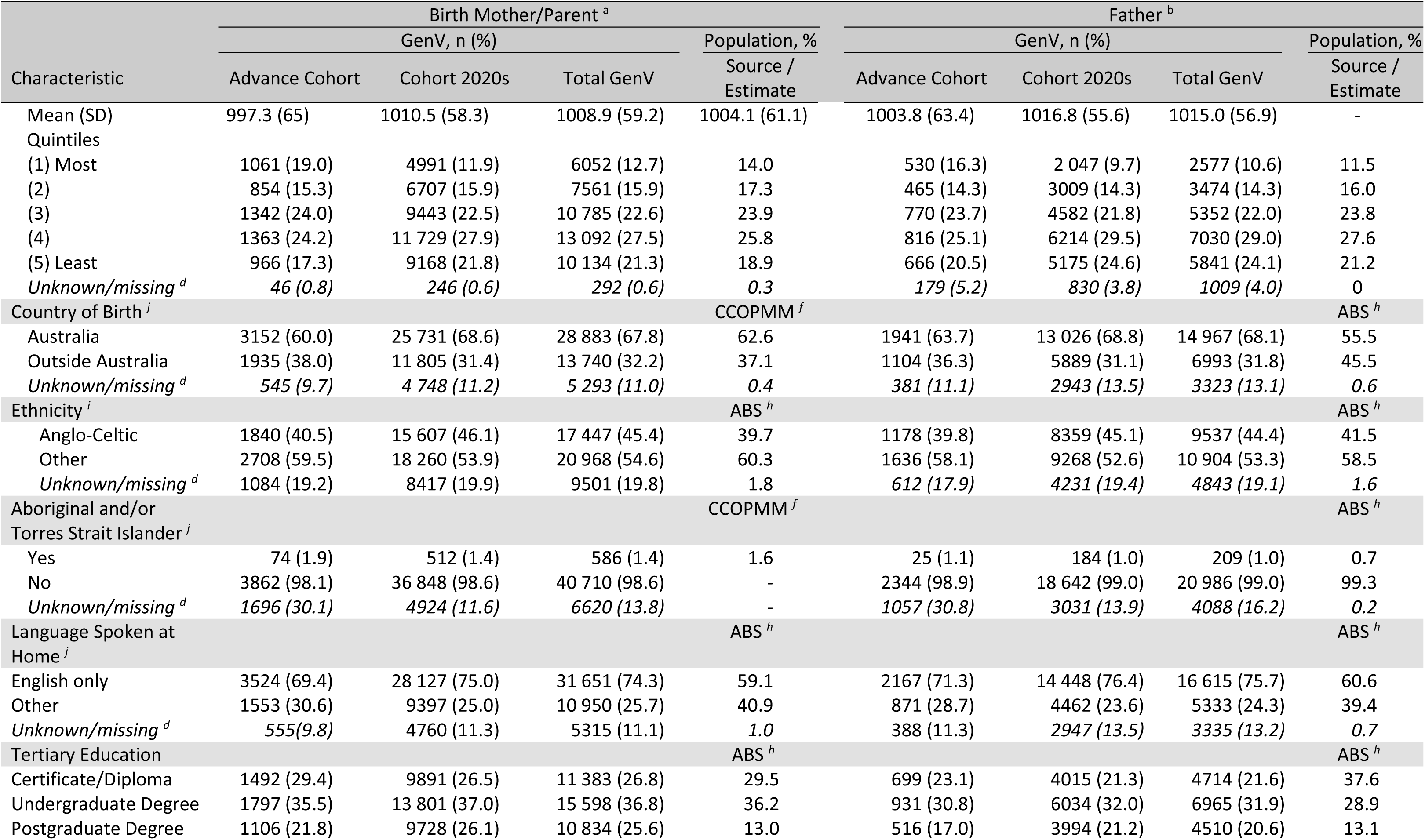
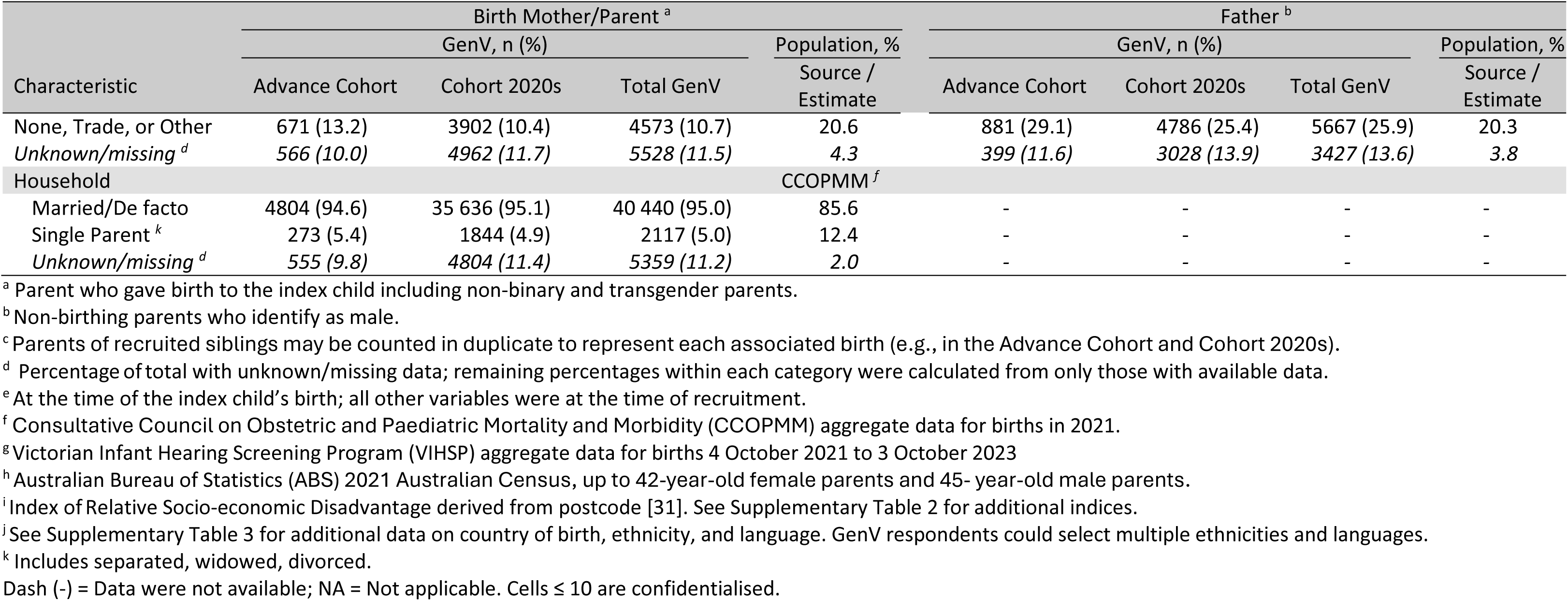
Birth mother/parent and father characteristics.

**Table 2.**
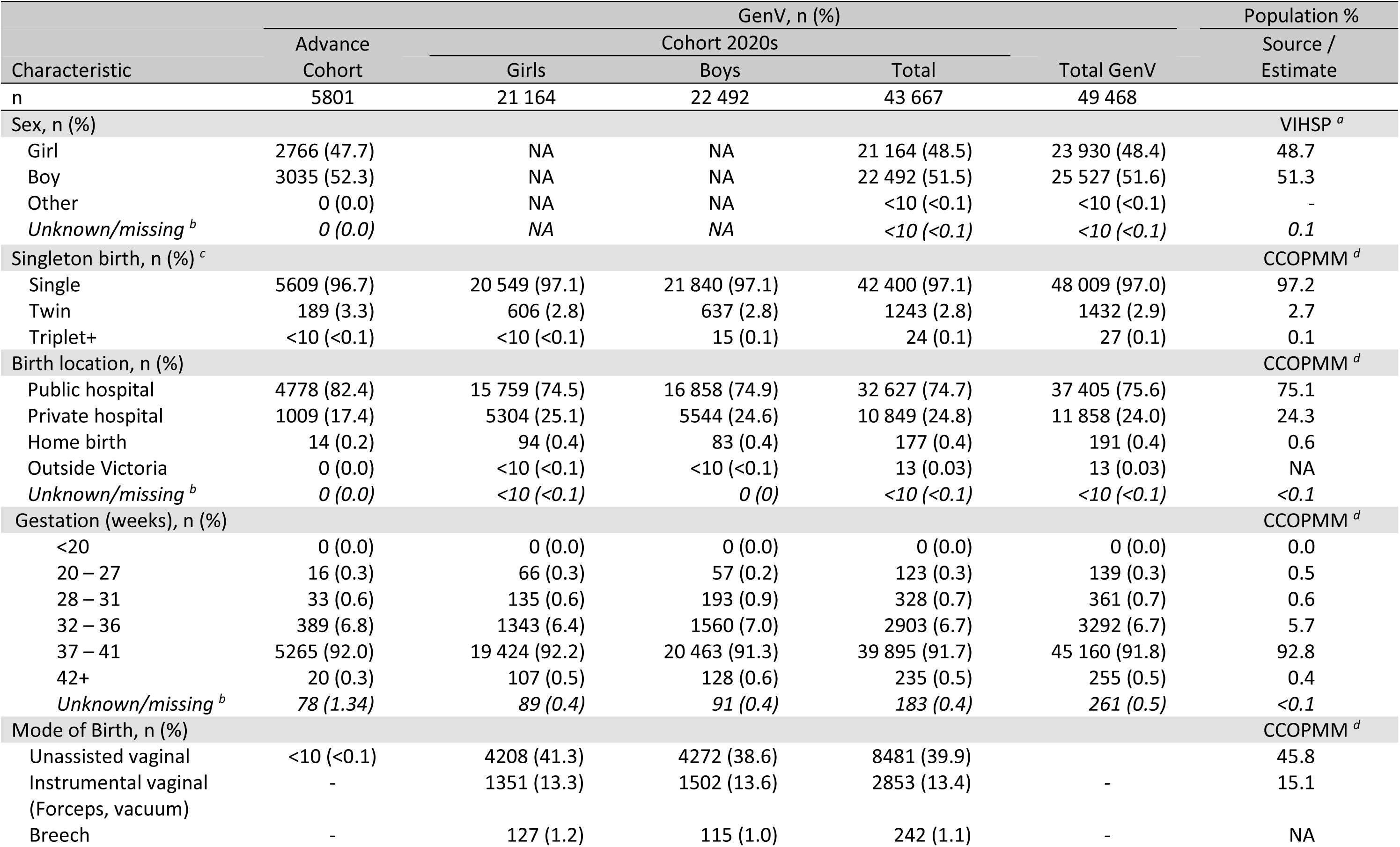
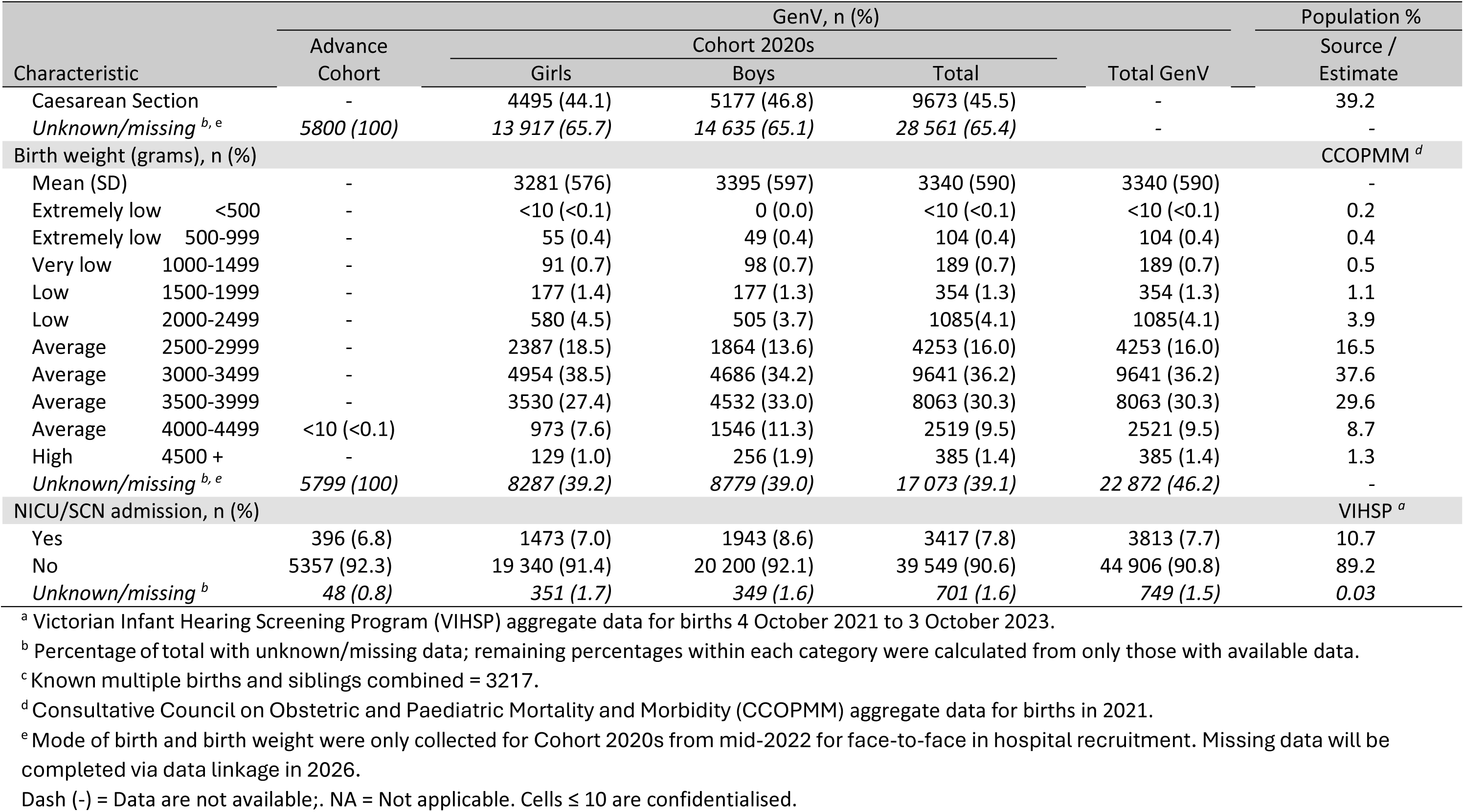
Child characteristics.

To demonstrate the representativeness of Cohort 2020s, we sourced the best currently available data for the Victorian population. (As the Advance Cohort was not Victoria-wide, we did not expect it to be representative.) Population source data were:

1. Consultative Council on Obstetric and Paediatric Mortality and Morbidity (CCOPMM): CCOPMM releases annual data for all births in Victoria. We used most recently available data tables for birth in 2021 [9].
2. Victorian Infant Hearing Screening Program (VIHSP): VIHSP reaches >99% of all Victorian births and provided GenV’s sampling frame, so provides the total proportion of responders. Aggregate data from all births recorded by VIHSP in the GenV eligible birth window were used for population comparisons.
3. Australian Bureau of Statistics (ABS) 2021 Australian Census: We used the ABS TableBuilder [10] to extract equivalent population data for Victorian resident mothers and fathers as reported by the Parent Indicator [11] (1=male parent and 2=female parent). While CCOPMM and VIHSP datasets were closely equivalent to GenV’s eligible birthing period, ABS Census included parents of children of all ages in the household and was therefore more likely to diverge from GenV; however, it was the only source available for fathers and some variables (e.g., education). To reduce capturing parents of older children, we restricted the maximum parental age to 42 years for mothers and 45 years for fathers, respectively (equivalent to mean age + 2 standard deviations of GenV birth mothers/parents and fathers, rounded up to the nearest integer).

GenV Cohort 2020s parents were representative of the population in regard to location of residence (e.g., metropolitan: 76.8% GenV birth mothers/parents v 76.0% VIHSP; 79.8% GenV fathers v 80.1% ABS), socio-economic disadvantage (e.g., most disadvantaged quintile: 11.9% GenV birth mothers/parents v 13.9% VIHSP), and identification as Aboriginal and/or Torres Strait Islander origin (1.4% GenV birth mothers/parents v 1.6% CCOPMM; 1.0% GenV fathers v 0.7% ABS). Among GenV birth mothers/parents, there was some under-representation of those who were younger (e.g., <30 years: 26.0% GenV v 30.5% CCOPMM) or single (5.0% GenV v 12.5% ABS). GenV birth mother/parents and fathers were more likely to have a postgraduate degree (26.1% GenV birth mothers/parents v 13.0% ABS; 21.2% GenV fathers v 13.1% ABS), and were somewhat less likely to be born outside Australia (31.4% GenV birth mothers/parents v 37.1% CCOPMM; 31.2% GenV fathers v 45.5% ABS), be of non-Anglo-Celtic background (53.9% GenV birth mothers/parents v 60.3% ABS; 52.6% GenV fathers v 58.5% ABS), or to speak languages other than English (25.0% GenV birth mothers/parents v 40.9% ABS; 23.6% GenV fathers v 39.4% ABS). Despite these differences, Supplementary Table 3 shows the wide cultural and linguistic diversity within the cohort.

GenV Cohort 2020s children resembled the population on sex (boys: 51.5% GenV v 51.3% VIHSP), multiple births (2.9% GenV v 2.8% CCOPMM), public hospital births (74.7% GenV v 75.1% CCOPMM), and birth weight (e.g., 3000g-3499g: 36.2% GenV v 37.6% CCOPMM). GenV included slightly fewer children admitted to special care nurseries or neonatal intensive care units (7.8% GenV v 10.7% VIHSP). Current data suggests GenV Cohort 2020s children had similar gestation (e.g., 37-41 weeks: 91.7% GenV v 92.8% CCOPMM) and slightly more caesarean section births (45.5% GenV v 39.2% CCOPMM) than the population; however, these figures (currently based on less than half the cohort) may shift with the addition of linked data in 2026.

## How often have they been followed up?

GenV comprises four core pre-defined elements: (1) recruitment (see above); (2) universal data linkages; (3) universal biosamples; and (4) major phenotypic waves every 5–6 years, starting in primary school. Optional elements include depth biosamples and the brief digital ‘GenV and Me’ surveys offered quarterly to age 12 months and then 6 monthly to age 5 years. Figure 2 shows the data and biosample coverage for the Cohort 2020s to date with further detail below and in our protocol [8].

**Figure 2:**
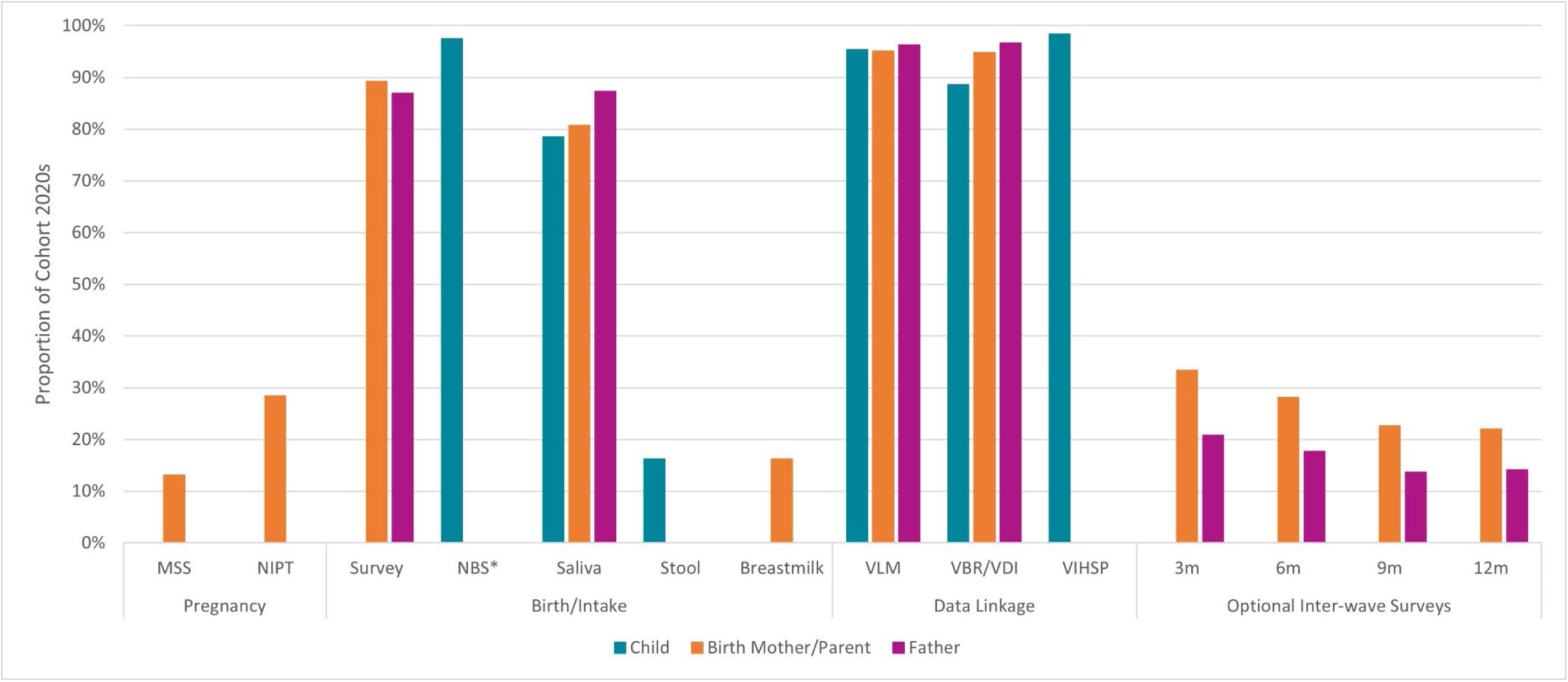
Data and biosamples available for Cohort 2020s *Pregnancy:* MSS = Maternal Serum Screen (n = 5562); NIPT = Non-invasive Prenatal Test (n = 12 010). Additional pregnancy biosamples collection is underway. *Birth/Intake:* Survey (n = 37 708 birth mothers/parents, 19 002 fathers); NBS = Newborn Bloodspot Screen matched records, final available samples may be lower (n = 42 543); saliva (n = 34 299 children, 34 131 birth mothers/parents, 19 087 fathers), day-7 stool (n = 7099) and breastmilk (n = 6568) collected October 2022-October 2023 for face-to-face recruited families only. *Data linkage:* VLM = Victorian Linkage Map, April 2024 (n = 41 631 children, 40 083 birth mothers/parents, 21 122 fathers); VBR/VDI = Victorian Births Registry and Victorian Death Index (n = 42 568 children, 40 518 birth mothers/parents, 21 017 fathers); VIHSP = Victorian Infant Hearing Screening Program (n = 42 972). Additional linked datasets are available (see Table 3) and underway. **Alt Text:** Bar graph depicting the percentage of the Cohort 2020s with available data and biosamples for birth mothers/parents, fathers, and children.

### Core data sources – Baseline survey, linkages, biosamples

GenV operates under a ‘low participant burden’ principle, avoiding duplication of routinely collected data. As of 1 May 2025, 89.2% of birth mothers/parents; 86.9% of fathers had completed the baseline survey. The first of GenV’s annual linkages via the Centre for Victorian Data Linkage (CVDL) connected to 15 state administrative datasets (Table 3), with 95.4% of participants matched to at least one dataset (indicated in Figure 2 by Victorian Linkage Map). The first geospatial datasets have been linked at address level for 93.7%, and 96.6% linkage has been achieved for births and deaths. Linkage will be expanded with annual refreshes including more state datasets and extensions to federal health and welfare datasets.

**Table 3.** GenV’s current external data sources (GenV linked and GenV extracted)

| **Dataset (Abbreviation)** | **Description** |
| --- | --- |
| **Source: Centre for Victorian Data Linkage** |  |
| Births, Deaths and Marriages (BDM) | *Contains official records of births, death and marriages registered in Victoria, including demographic and parental information.* |
| Child Protection Services Dataset (CP) | *Tracks reports, investigations, and outcomes related to child protection and welfare.* |
| Community Health Services (CHMDS) | *Includes service delivery data from community health centres across Victoria.* |
| Family Services - Cradle 2 Kinder (IRIS C2K) | *Provides data on early intervention and support services (pre-birth support, intensive and longer-term intervention and case work support) for vulnerable families until child reaches 4 years of age.* |
| Family Services (IRIS FS) | *Tracks broader family support services aimed at improving child and family wellbeing.* |
| Family Violence (IRIS FV) | *Contains information on services provided to individuals affected by family violence.* |
| Mental Health Community Support Services (MHCSS) | *Documents support services for individuals with mental health conditions living in the community.* |
| Public Health Event Surveillance System (PHESS) | *Monitors notifiable diseases and public health incidents.* |
| Transmission Response Epidemiology Victoria Information System (TREVI) | *Tracks infectious disease transmission and public health responses.* |
| Victorian Admitted Episodes Collection (VAED) | *Contains comprehensive data on hospital admissions, diagnoses, and procedures.* |
| Victorian Alcohol and Drug Collection (VADC) | *Records treatment episodes for alcohol and drug-related issues.* |
| Victorian Cost Data Collection (VCDC) | *Provides cost and activity data for public hospital services. Now, incorporated within Admitted episodes, emergency department and non-admitted episodes* |
| Victorian Death Index - Cause of Death | *Contains causes of death from official death registrations.* |
| Victorian Emergency Minimum Dataset (VEMD) | *Captures administrative and clinical data on emergency department presentations and outcomes at public hospitals.* |
| Victorian Integrated Non-Admitted Health (VINAH) | *Records service use and outcomes for non-admitted patients in public health services.* |
| **Source: Royal Children's Hospital** |  |
| Victorian Infant Hearing Screening Program (VIHSP) | *Captures outcomes of infant hearing screenings, follow-up diagnostics, and associated risk factors for hearing impairment.* |
| **Source: Various birthing hospitals <sup>a</sup>** |  |
| Hospital administrative datasets | *Includes datasets prepared for VAED, VEMD, VINAH* |
| Birthing Outcomes System (BOS) | *Documents pregnancy, birth, neonatal, postpartum, and domiciliary care outcomes. VPDC minimum data included.* |

| Dataset (Abbreviation) | Description |
| --- | --- |
| Selected data categories ('modules') | *Sourced from Electronic Medical Records/paper-based records; example include prescribing, allergies and sensitivities, clinical problem lists, care parameters, discharge documents / discharge summary, vital signs.* |
| Reports and results | *Examples include imaging reports and pathology results.* |

<sup>a</sup> In progress with multiple sites.
Further metadata available at https://metadata.phrn.org.au/ and https://vahi.vic.gov.au/ourwork/data-linkage/apply

Some collections are completed once only at specific life stages. For example, infant hearing screening data have been transferred to GenV (98.4% matched), and statewide extraction of detailed birthing hospital medical records is in progress.

Saliva samples were collected postnatally for 80.7% of birth mothers/parents, 87.3% of fathers, and 78.5% of children. Custodianship transfer is in progress for newborn bloodspot samples (97.4% records matched; final sample availability may be lower). GenV’s partnerships have also supported Victorian pathology services to retain residual antenatal and perinatal biosamples for transfer to GenV post-consent, with matching and transfer under way. Consent to genetic research was given for 89.0% of birth mothers/parents, 91.2% of fathers, and 88.0% of children.

### Optional sources – Depth biosamples, GenV and Me

‘Depth’ biosamples include a sub-cohort of Day 7 infant stool (16.3%) and breastmilk (15.5%), with follow-up stool samples at age 2 years. *’GenV and Me*’ surveys to age 12 months were completed by 22.0%-33.3% birth mothers/parents and 13.7%-20.8% fathers.

### Integrated Studies

Families may join embedded collaborative studies, including randomised trials, natural experiments and deep subcohorts. See [12–14] for details.

### Withdrawals

As of 1 May 2025, 678 (0.6%) of Cohort 2020s participants had withdrawn from all further data collection.

## What has been measured?

Measures align with GenV’s outcomes hierarchy [15], life course framework [16], review of core outcome sets [17], and Global Burden of Disease priorities for high-income countries.

### Baseline survey

Completed immediately or shortly after recruitment, this brief survey collects key demographic, physical and mental health, and pregnancy/labour experience data.

### Linked data

Participants consent to linkage across health, education, social and neighbourhood domains. GenV’s website [18] lists all data linkage sources, updated annually. Initial state datasets cover hospital/emergency visits, immunisation, infectious diseases and a range of community services. See Table 3 for a list of currently linked datasets. Planned national linkages include to Australia’s PLIDA (Person-Level Integrated Data Asset), NDDA (National Disability Data Asset) and NHDH (National Health Data Hub).

Hospital records include curated datasets for state reporting (e.g., admissions, emergency presentations, perinatal indicators) and uncurated data (e.g., inpatient prescribing, pathology, imaging) not captured in administrative collections. VIHSP data provide results of universal newborn hearing screens and follow-up audiology diagnoses.

### Biosamples

Our published protocol [8] shows GenV’s wide-ranging but small-volume biosamples and their potential uses and value. First bioassays are likely to be made available by 2029. Biosamples include residual clinical samples (e.g., newborn bloodspots, maternal antenatal samples) and participant-collected GenV samples (e.g., saliva, infant stool, breastmilk).

### GenV and Me

collects brief digital surveys, photos and videos. Metadata can be browsed on the MCRI LifeCourse Initiative website https://lifecourse.melbournechildrens.com/cohorts/genv/.

### Phenotypic waves

Major waves around ages 6, 11, and 16, funding permitting, will include in-school and remote components with standardised, objective phenotyping prioritised by disease burden. GenV welcomes collaboration to inform content for the upcoming Early School Wave, especially for scalable, innovative measures.

## What has it found?

GenV’s early publications have focused on the ways in which very large population cohorts can optimise relevance, timeliness, utility and future impact to promote wellbeing and reduce burden of major current and future problems facing today’s children and pre-midlife adults. Publications have synthesised Core Outcome Sets to prioritise the measures consistently of highest value to families, clinicians and services across multiple conditions and across the life course [17]. Two Discrete Choice Experiments with over 3,000 GenV parents found that, while all health areas related to the top ten causes of global burden of disease were seen as important, parents’ top priorities for their children were anxiety/depression, long-term heart health, autism and asthma [19]. A synthesis of life course phenotypic trajectories revealed major gaps for many noncommunicable diseases [20], while a parallel review of current large children’s cohorts worldwide showed content choices that are likely to leave these gaps unfilled - reinforcing GenV’s unique potential to do so [21].

GenV has also advanced methods to integrate registries, trials and digital governance into whole-population cohorts, including co-participation models enabling embedded trials and health services research [12–14, 22]. For example, funded whole-population collaborations are already testing cost effectiveness of new screening workflows for congenital cytomegalovirus and for newborn conditions not in current newborn bloodspot programs, with outcomes expected by 2028. See GenV’s peer-reviewed and working paper publications on our website [18].

## What are the main strengths and weaknesses?

GenV is the only mega-birth cohort recruited throughout the systemic shocks of the early 2020s (pandemic, geopolitical, economic, climate and environment), positioning it for solutions to contemporary problems and to enable developmental origins of health and disease (DOHaD) discoveries, such as life course impacts of viruses on brain health and cognition [23, 24].

With nearly 50 000 children and 74 000 parents from across an entire Australian state the size of the UK, GenV is notable for its high uptake (29.7% of an entire birthing population) and inclusive, multilingual cohort (see Supplementary Table 3). Whilst not applicable to remote Australian communities, GenV includes a sizable number of parents who identified as First Nations and what we believe is Australia’s largest known rural/regional birth cohort. It also includes groups under-represented in European and American mega-cohorts, such as those with southern, south-eastern, eastern and western Asian heritages [25–29]. Ongoing recruitment is expected to further increase representation, including new migrants with children born in GenV’s eligible birth window. As such, the Cohort 2020s’ profile will evolve over time, and we expect to publish profile updates.

Of note, in contrast to the primarily maternal cohorts of the past, GenV offers independent participation to all parents/guardians, regardless of gender or birth relationship. This will enable research into the ‘missing middle’ of adult life – pre-midlife health and ageing [30]. However, we acknowledge that the parent cohort excludes adults who do not have children.

Our temporospatial, low-burden, universal data matrix spans major contributors to health and disease from cell to society, requiring trade-offs between brevity and breadth vs depth. Biosamples require prioritisation and small-volume assays. Optional GenV and Me surveys offer rich data but with lower response rates. Our planned 5-yearly universal phenomic assessments, tailored to concerns of parents and educators in Victorian schools, will likely enable higher-response phenotypic, survey, and biosample collection. However, sustainability remains a challenge in the absence of dedicated cohort funding pathways in Australia.

GenV has already demonstrated success in matching universal ante/perinatal biosamples, data linkage, and experiential/phenotypic data collection, and is building cloud infrastructure to support Open Science. Over AUD$30 million in competitive research funding has been secured, with >150 collaborators. GenV’s proposed Intervention Hub – to serve as a population registry enabling trials at public health scale – is, to our knowledge, globally unique.

## Can I get hold of the data? Where can I find out more?

Collaborative proposals aligning with GenV’s guiding principles [8] are welcomed now via www.genv.org.au. End-user access to early data and biosamples applications is expected from late 2026. GenV is registered at ClinicalTrials.gov: NCT05394363.

## Ethics approval

GenV is approved by The Royal Children’s Hospital Melbourne Human Research Ethics Committee (HREC/51302/RCHM-2019; local reference 2019.011). Parents/guardians provide written informed consent for their own and their child’s participation.

## Supplementary data

Supplementary data are available at IJE online.

## Author contributions

MW, SG, RS and WS were central to the conception of GenV. EKH, WS, AG, SAC, TF, JM, NS, MB, ZP, JS, NZ, SG, RS, and MW contributed to the design and implementation of GenV. EKH, AF and DS undertook the analyses. EKH and MW drafted the paper and all authors reviewed and revised drafts. All authors have approved the submitted version.

## Use of artificial intelligence (AI) tools

EKH used Copilot to assist with writing description of datasets in Table 3, which were verified by JM and another team member. DS wrote original analytic code then used ChatGPT for optimisation and refactoring of some code. MW used ChatGPT for minor revisions to flow of text.

## Funding

This work was supported by the Paul Ramsay Foundation, the Victorian State Government, The Royal Children’s Hospital Foundation, the Murdoch Children’s Research institute, the National Health and Medical Research Council and the Medical Research Future Fund (NCRI000077; MRFRDIII000002). Research at the MCRI is supported by the Victorian Government’s Operational Infrastructure Support Program. MW is supported by Australian National Health and Medical Research Council (NHMRC) Investigator Grant (2035040). SG is supported by NHMRC Investigator Grant (2026263). GenV is led from MCRI, in partnership with The Royal Children’s Hospital and University of Melbourne.

## Data Availability

All data produced in the present study are available upon reasonable request to the authors.

## Acknowledgements

We thank: The GenV Investigator Committee (in alphabetical order and inclusive of past and present members: Dino Asproloupos, Katie Allen, James Boyd, David Burgner, Jim Buttery, John Carlin, Jeanie Cheong, Nigel Curtis, Ben Edwards, Harriet Hiscock, Tu’uhevaha Kaitu’u-Lino, Suzanne Mavoa, Fiona Mensah, Kathryn North, Anne-Louise Ponsonby, Elenor Williams, Katrina Williams, Valerie Sung, Mary Wlodek); the GenV Site Principal Investigators and Liaisons; Sheena Reilly; Victorian Infant Hearing Screening Program staff; members of the GenV Working Groups and Steering Committee; the staff and patients at all our recruitment sites; and all the GenV staff past and present and the supporting departments of the MCRI and RCH. GenV acknowledges the Traditional Custodians of lands on which we work across Victoria. We pay our respects to their Elders past, present and emerging.

## Conflict of interest

None declared.

## Notes

### Competing Interest Statement

The authors have declared no competing interest.

### Author Declarations

The Human Research Ethics Committee of The Royal Children's Hospital Melbourne gave ethical approval for this work.

