## Supplemental Tables for "Cohort Profile: Generation Victoria (GenV)"

**SUPPLEMENTARY TABLE 1 Combined GenV parent cohort (all mothers, fathers, other parents and guardians)**

| Characteristic | Advance Cohort | Cohort 2020s | Total GenV |
|---|---|---|---|
| n | 9081 | 64 428 | 73 509 |
| Relationship to index child, n (%) [a] | | | |
| Birthing parent/guardian | | | |
| Guardian (mother/father/parent) | 5632 (62.0) | 42 284 (65.6) | 47 916 (65.2) |
| Surrogate | <10 (<0.1) | <10 (<0.1) | <10 (<0.1) |
| Non-birthing parent/guardian | | | |
| Father (incl biological, step, intended) | 3426 (37.7) | 21 857 (33.9) | 25 283 (34.4) |
| Mother (including biological, intended) | 18 (0.2) | 245 (0.4) | 263 (0.4) |
| Grandparent | <10 (<0.1) | 14 (<0.1) | 16 (<0.1) |
| Other (eg aunt) | <10 (<0.1) | 28 (<0.1) | 31 (<0.1) |
| Gender, n (%) | | | |
| Female/Woman | 5653 (62.2) | 42 491 (65.9) | 48 144 (65.5) |
| Male/Man | 3427 (37.7) | 21 885 (34.0) | 25 312 (34.4) |
| Other | <10 (0.01) | 52 (0.08) | 53 (0.07) |
| *Unknown/missing* [b] | *0 (0.0)* | *0 (0.0)* | *0 (0.0)* |
| Age (years) at birth of child [c] | | | |
| M (SD) | 32.3 (5.1) | 32.9 (5.0) | 32.8 (5.0) |
| Median | 32 | 33 | 33 |
| Range | 15-66 | 14-72 | 14-72 |
| Categorical, n (%) | | | |
| <20 | 66 (0.7) | 327 (0.5) | 393 (0.5) |
| 20-24 | 511 (5.6) | 2782 (4.3) | 3293 (4.5) |
| 25-29 | 1915 (21.2) | 11 838 (18.4) | 13 753 (18.7) |
| 30-34 | 3707 (41.0) | 26 189 (40.7) | 29 896 (40.7) |
| 35-39 | 2219 (24.5) | 17 864 (27.7) | 20 083 (27.3) |
| 40-44 | 511 (5.6) | 4523 (7.0) | 5034 (6.9) |
| 45+ | 121 (1.3) | 852 (1.3) | 973 (1.3) |
| *Unknown/missing* [b] | *31 (0.3)* | *54 (0.1)* | *85 (0.1)* |

| Characteristic | Advance Cohort | Cohort 2020s | Total GenV |
|---|---|---|---|
| Location of residence, n (%) | | | |
| Major City | 7154 (80.8) | 49 256 (77.8) | 56 410 (78.1) |
| Inner Regional | 1505 (17.0) | 12 117 (19.1) | 13 622 (18.9) |
| Outer Regional | 194 (2.2) | 1945 (3.1) | 2139 (3.0) |
| Remote | 0 (0.0) | 12 (<0.1) | 12 (<0.1) |
| Very Remote | 0 (0.0) | <10 (<0.1) | <10 (<0.1) |
| *Unknown/missing* [b] | *228 (2.5)* | *1095 (1.7)* | *1323 (1.8)* |
| Socioeconomic Status, n (%) | | | |
| Relative Socio-economic Disadvantage [d] | | | |
| Mean (SD) | 999.7 (64.5) | 1012.6 (57.4) | 1011.0 (58.5) |
| Quintiles | | | |
| (1) Most disadvantage | 1596 (18.0) | 7067 (11.2) | 8663 (12.0) |
| (2) | 1323 (14.9) | 9768 (15.42) | 11 091 (15.4) |
| (3) | 2114 (23.9) | 14 071 (22.2) | 16 185 (22.4) |
| (4) | 2184 (24.7) | 18 012 (28.4) | 20 196 (28.0) |
| (5) Least disadvantage | 1636 (18.5) | 14 415 (22.8) | 16 051 (22.2) |
| *Unknown/missing* [b] | *228 (2.5)* | *1095 (1.7)* | *1323 (1.8)* |
| Country of Birth [e] | | | |
| Australia | 5110 (62.6) | 38 960 (68.7) | 44 070 (67.9) |
| Outside Australia | 3044 (37.38) | 17 747 (31.3) | 20 791 (32.1) |
| Non-English speaking country | 2482 (30.4) | 13 788 (24.3) | 16 270 (25.1) |
| *Unknown/missing* [b] | *927 (10.2)* | *7722 (12.0)* | *8649 (11.8)* |
| Ethnicity [e] | | | |
| Anglo-Celtic | 3031 (41.0) | 24 109 (46.6) | 27 142 (45.9) |
| Other | 4353 (58.9) | 27 623 (53.4) | 31 976 (54.1) |
| *Unknown/missing* [b] | 1697 (18.7) | 12 697 (19.7) | 14 394 (19.6) |
| Aboriginal and/or Torres Strait Islander [e] | | | |
| Yes | 100 (1.6) | 699 (1.2) | 799 (1.3) |
| No | 6224 (98.4) | 55 742 (98.8) | 61 966 (98.7) |
| *Unknown/missing* [b] | *2757 (30.3)* | *7988 (12.4)* | *10 745 (14.6)* |

| Characteristic | Advance Cohort | Cohort 2020s | Total GenV |
|---|---|---|---|
| Language Spoken at Home [e] | | | |
| English only | 5712 (70.2) | 42 805 (75.5) | 48 517 (74.8) |
| Other | 2425 (29.8) | 13 885 (24.5) | 16 310 (25.2) |
| *Unknown/missing* [b] | *944 (10.4)* | *7738 (12.0)* | *8682 (11.8)* |
| Tertiary Education | | | |
| Certificate/Diploma | 2195 (27.0) | 13 977 (24.8) | 16 172 (25.1) |
| Undergraduate Degree | 2733 (33.7) | 19 922 (35.3) | 22 655 (35.1) |
| Postgraduate Degree | 1629 (20.1) | 13 785 (24.4) | 15 414 (23.9) |
| None, Trade, or Other | 1558 (19.2) | 8723 (15.4) | 10 281 (16.0) |
| *Unknown/missing* [b] | *967 (10.6)* | *8020 (12.4)* | *8987 (12.2)* |
| Household | | | |
| Married/De facto | 4806 (94.6) | 35 636 (95.0) | 40 442 (95.0) |
| Single Parent [f] | 274 (5.4) | 1844 (5.0) | 2118 (5.0) |
| *Unknown/missing* [b] | *4001 (44.1)* | *26 949 (41.8)* | *30 950 (42.1)* |

[a] As reported by the parent/guardian at time of recruitment; Surrogate refers to the gestational carrier and intended refers to the person who will be parent/guardian (also called commissioning parent). Parents of recruited siblings may be counted in duplicate to represent each associated birth (e.g., in the Advance Cohort and Cohort 2020s).

[b] Percentage of total with unknown/missing data; remaining percentages within each category were calculated from only those with available data.

[c] At the time of the index child's birth; all other variables are at the time of recruitment.

[d] Index of Relative Socio-economic Disadvantage derived from postcode [31]. See Supplementary Table 2 for additional indices.

[e] See Supplementary Table 3 for additional data on country of birth, ethnicity and language. GenV respondents could select multiple ethnicities and languages.

[f] Includes separated, widowed, divorced.

Cells ≤ 10 are confidentialised.

**SUPPLEMENTARY TABLE 2 Socio-economic status quintiles for GenV birth mothers/parents (compared to population) and GenV fathers [a]**

| Characteristic | Birth Mother/Parent [b] GenV, n (%) Advance Cohort | Cohort 2020s | Total GenV | Population, % Source / Estimate | Father [c] GenV, n (%) Advance Cohort | Cohort 2020s | Total GenV |
|---|---|---|---|---|---|---|---|
| n | 5632 | 42 284 | 47 916 | | 3426 | 21 857 | 25 283 |
| Index of Relative Socio-economic Disadvantage (IRSD) | | | | VIHSP [d] | | | |
| Mean (SD) | 997.3 (65) | 1010.5 (58.3) | 1008.9 (59.2) | | 1003.8 (63.4) | 1016.8 (55.6) | 1015.0 (56.9) |
| Quintiles | | | | | | | |
| (1) Most Disadvantaged | 1061 (19.0) | 4991 (11.9) | 6052 (12.7) | 13.9 | 530 (16.3) | 2047 (9.7) | 2577 (10.6) |
| (2) | 854 (15.3) | 6707 (15.9) | 7561 (15.9) | 17.3 | 465 (14.3) | 3009 (14.3) | 3474 (14.3) |
| (3) | 1342 (24.0) | 9443 (22.5) | 10 785 (22.6) | 23.8 | 770 (23.7) | 4582 (21.8) | 5352 (22.0) |
| (4) | 1363 (24.2) | 11 729 (27.9) | 13 092 (27.5) | 25.6 | 816 (25.1) | 6214 (29.5) | 7030 (29.0) |
| (5) Least Disadvantaged | 966 (17.3) | 9168 (21.8) | 10 134 (21.3) | 19.0 | 666 (20.5) | 5175 (24.6) | 5841 (24.1) |
| Index of Relative Socio-economic Advantage and Disadvantage (IRSAD) | | | | | | | |
| Mean (SD) | 993.7 (66.4) | 1008.0 (66.1) | 1006.3 (66.3) | | 1001.6 (66.8) | 1016.0 (65.4) | 1013.9 (65.8) |
| Quintiles | | | | | | | |
| (1) Most Disadvantaged | 957 (17.1) | 4735 (11.3) | 5692 (11.9) | 13.0 | 479 (14.7) | 1954 (9.3) | 2433 (10.0) |
| (2) | 655 (11.7) | 5266 (12.5) | 5921 (12.4) | 13.4 | 336 (10.3) | 2257 (10.7) | 2593 (10.7) |
| (3) | 1464 (26.2) | 9586 (22.8) | 11 050 (23.2) | 24.2 | 844 (26.0) | 4615 (21.9) | 5459 (22.5) |
| (4) | 1438 (25.7) | 11 279 (26.8) | 12 717 (26.7) | 25.1 | 834 (25.7) | 5778 (27.5) | 6612 (27.2) |
| (5) Least Disadvantaged | 1072 (19.2) | 11 172 (26.6) | 12 244 (25.7) | 23.9 | 754 (23.2) | 6423 (30.5) | 7177 (29.6) |
| Index of Economic Resources (IER) | | | | | | | |
| Mean (SD) | 1007.1 (55.3) | 1008.9 (54.2) | 1008.7 (54.3) | | 1009.3 (55.1) | 1010.6 (53.5) | 1010.4 (53.7) |
| Quintiles | | | | | | | |
| (1) Most Disadvantaged | 934 (16.7) | 6837 (16.3) | 7771 (16.3) | 17.5 | 520 (16.0) | 3211 (15.3) | 3731 (15.4) |
| (2) | 832 (14.9) | 5877 (14.0) | 6709 (14.1) | 14.2 | 463 (14.3) | 2844 (13.5) | 3307 (13.6) |
| (3) | 924 (16.5) | 7487 (17.8) | 8411 (17.7) | 16.3 | 541 (16.7) | 3877 (18.4) | 4418 (18.2) |
| (4) | 1825 (32.7) | 13 037 (31.0) | 14 862 (31.2) | 32.1 | 1060 (32.6) | 6517 (31.0) | 7577 (31.2) |
| (5) Least Disadvantaged | 1071 (19.2) | 8800 (20.9) | 9871 (20.7) | 19.3 | 663 (20.4) | 4578 (21.8) | 5241 (21.6) |

| Characteristic | Birth Mother/Parent [b] GenV, n (%) Advance Cohort | Birth Mother/Parent [b] GenV, n (%) Cohort 2020s | Birth Mother/Parent [b] GenV, n (%) Total GenV | Birth Mother/Parent [b] Population, % Source / Estimate | Father [c] GenV, n (%) Advance Cohort | Father [c] GenV, n (%) Cohort 2020s | Father [c] GenV, n (%) Total GenV |
|---|---|---|---|---|---|---|---|
| Index of Education and Occupation (IEO) | | | | | | | |
| Mean (SD) | 996.8 (69.0) | 1013.4 (72.8) | 1011.4 (72.6) | | 1005.3 (70.8) | 1022.1 (73.3) | 1019.8 (73.2) |
| Quintiles | | | | | | | |
| (1) Most Disadvantaged | 479 (8.6) | 2442 (5.8) | 2921 (6.1) | 6.9 | 225 (6.9) | 944 (4.5) | 1169 (4.8) |
| (2) | 900 (16.1) | 5628 (13.4) | 6528 (13.7) | 14.3 | 484 (14.9) | 2420 (11.5) | 2904 (12.0) |
| (3) | 1678 (30.0) | 10 240 (24.4) | 11 918 (25.0) | 25.3 | 906 (27.9) | 4789 (22.8) | 5695 (23.5) |
| (4) | 1403 (12.1) | 11 939 (28.4) | 13 342 (28.0) | 27.6 | 844 (26.0) | 6085 (28.9) | 6929 (28.5) |
| (5) Least Disadvantaged | 1126 (20.2) | 11 789 (28.0) | 12 915 (27.1) | 25.4 | 788 (24.3) | 6789 (32.3) | 7577 (31.2) |
| *Unknown/Missing [e]* | *46 (0.8)* | *246 (0.6)* | *292 (0.6)* | *0.5* | *179 (5.2)* | *830 (3.8)* | *1009 (4.0)* |

[a] Socioeconomic Indexes for Areas [31] derived from postcode at the time of recruitment.

[b] Parent who gave birth to the index child including non-binary and transgender parents.

[c] Non-birthing parents who identify as male.

[d] Victorian Infant Hearing Screening Program (VIHSP) aggregate data for births 4 October 2021 to 3 October 2023.

[e] Percentage of total with unknown/missing data; remaining percentages within each category were calculated from only those with available data.

**SUPPLEMENTARY TABLE 3 Cultural and linguistic diversity of GenV birth mothers/parents and fathers**

| | Birth Mother/Parent [a] | | | | Father [b] | | | |
|---|---|---|---|---|---|---|---|---|
| | GenV, n (%) | | | Population, % | GenV, n (%) | | | Population, % |
| Characteristic | Advance Cohort | Cohort 2020s | Total GenV | Source / Estimate | Advance Cohort | Cohort 2020s | Total GenV | Source / Estimate |
| n | 5 632 | 42 284 | 47 916 | | 3 426 | 21 857 | 25 283 | |
| Region/Country of Birth [c] | | | | CCOPMM [d] | | | | ABS [e] |
| Oceania/Antarctica | 3316 (63.2) | 26 651 (71.1) | 29 967 (70.3) | 65.2 | 2021 (66.3) | 13 464 (71.1) | 15 485 (70.5) | 58.3 |
| Australia | 3152 (60.0) | 25 731 (68.6) | 28 883 (67.8) | 62.6 | 1941 (63.7) | 13 026 (68.8) | 14 967 (68.1) | 55.5 |
| Southern & Central Asia | 727 (17.3) | 3753 (10.0) | 4 480 (10.5) | 13.3 | 481 (15.8) | 2069 (10.9) | 2550 (11.6) | 18.2 |
| India | 482 (9.5) | 2380 (6.3) | 2862 (6.7) | 8.3 | 319 (10.5) | 1 339 (7.1) | 1 658 (7.5) | 12.6 |
| Sri Lanka | 96 (1.9) | 551 (1.5) | 647 (1.5) | 1.4 | 71 (2.3) | 329 (1.7) | 400 (1.8) | 1.9 |
| Pakistan | 62 (1.2) | 306 (0.8) | 368 (0.9) | 1.4 | 36 (1.2) | 149 (0.8) | 185 (0.8) | 1.4 |
| Afghanistan | 30 (0.6) | 158 (0.4) | 188 (0.4) | 1.0 | 20 (0.7) | 70 (0.4) | 90 (0.4) | 1.0 |
| Nepal | 39 (0.8) | 254 (0.7) | 293 (0.7) | 0.8 | 26 (0.8) | 123 (0.6) | 149 (0.7) | 0.7 |
| South-East Asia | 388 (7.6) | 2121 (5.6) | 2509 (5.9) | 6.0 | 173 (5.7) | 765 (4.0) | 938 (4.3) | 6.1 |
| Vietnam | 130 (2.5) | 418 (1.1) | 548 (1.3) | 1.6 | 54 (1.8) | 145 (0.8) | 199 (0.9) | 1.6 |
| Philippines | 111 (2.2) | 664 (1.8) | 775 (1.8) | 1.4 | 53 (1.7) | 249 (1.3) | 302 (1.4) | 1.5 |
| Malaysia | 56 (1.1) | 471 (1.2) | 527 (1.2) | 1.0 | 31 (1.0) | 193 (1.0) | 224 (1.0) | 1.1 |
| North-East Asia | 132 (2.6) | 1408 (3.7) | 1540 (3.6) | 4.0 | 66 (2.2) | 560 (3.0) | 626 (2.8) | 5.2 |
| China | 90 (1.8) | 1010 (2.7) | 1100 (2.6) | 2.7 | 48 (1.6) | 409 (2.2) | 457(2.1) | 4.0 |
| North African/Middle East | 108 (2.12) | 537 (1.4) | 645 (1.5) | 3.2 | 44 (1.4) | 247 (1.3) | 291 (1.3) | 3.4 |
| Iraq | 20 (0.4) | 74 (0.2) | 94 (0.2) | 0.7 | 10 (0.3) | 25 (0.1) | 35 (0.2) | 0.7 |
| North-West Europe | 162 (3.2) | 1364 (3.6) | 1526 (3.6) | 2.8 | 126 (4.2) | 931 (4.9) | 1057 (4.8) | 3.6 |
| Sub-Saharan Africa | 91 (1.8) | 563 (1.5) | 654 (1.5) | 2.1 | 51 (1.7) | 293 (1.5) | 344 (1.6) | 1.9 |
| Southern/Eastern Europe | 74 (1.4) | 444 (1.2) | 515 (1.2) | 1.6 | 38 (1.2) | 212 (1.1) | 250 (1.1) | 2.0 |
| Americas | 79 (1.5) | 670 (1.8) | 749 (1.8) | 1.5 | 44 (1.4) | 358 (1.9) | 402 (1.8) | 1.3 |
| *Unknown/missing [f]* | *545 (9.7)* | *4 748 (11.2)* | *5 293 (11.0)* | *0.4* | *381 (11.1)* | *2943 (13.5)* | *3323 (13.1)* | *0.6* |
| Country of Birth by English-speaking status | | | | CCOPMM [d] | | | | ABS [e] |
| Born in English-speaking country | 3500 (68.8) | 28 190 (75.1) | 31 690 (74.3) | 68.1 | 2152 (70.7) | 14 497 (76.6) | 16 649 (75.8) | 61.7 |
| Born in non-English-speaking country | 1587 (31.2) | 9346 (24.9) | 10 933 (25.6) | 31.9 | 893 (29.3) | 4418 (23.4) | 5311 (24.2) | 38.3 |
| *Unknown/Missing [f]* | *545 (9.7)* | *4 748 (11.2)* | *5 293 (11.0)* | *0.4* | *381 (11.1)* | *2943 (13.5)* | *3323 (13.1)* | *0.6* |

| | Birth Mother/Parent [a] | | | | Father [b] | | | |
|---|---|---|---|---|---|---|---|---|
| | GenV, n (%) | | | Population, % | GenV, n (%) | | | Population, % |
| Characteristic | Advance Cohort | Cohort 2020s | Total GenV | Source / Estimate | Advance Cohort | Cohort 2020s | Total GenV | Source / Estimate |
| Ethnic Background [g] | | | | ABS [e] | | | | ABS [e] |
| Aboriginal and/or Torres Strait Islander | 69 (1.28) | 510 (1.2) | 579 (1.2) | 1.2 | 24 (0.7) | 182 (0.9) | 206 (0.8) | 0.9 |
| African | 117 (2.2) | 617 (1.5) | 734 (1.6) | 1.7 | 44 (1.3) | 259 (1.2) | 303 (1.2) | 1.8 |
| Anglo-Celtic | 2176 (40.3) | 19 046 (46.5) | 21 222 (45.9) | 39.7 | 1430 (43.2) | 10 344 (48.9) | 11 774 (48.2) | 41.5 |
| European - Southern | 481 (8.92) | 3598 (8.8) | 4079 (8.8) | 10.6 | 302 (9.1) | 1885 (8.9) | 2187 (8.9) | 11.4 |
| European - Northern/ Western/ Eastern | 463 (8.6) | 4044 (9.9) | 4507 (9.7) | 8.4 | 369 (11.1) | 2460 (11.6) | 2829 (11.6) | 8.4 |
| East Asian | 225 (4.2) | 2243 (5.5) | 2468 (5.3) | 7.2 | 133 (4.0) | 976 (4.6) | 1109 (4.5) | 8.0 |
| Southeast Asian | 451 (8.4) | 2431 (5.9) | 2882 (6.2) | 9 | 204 (6.2) | 977 (4.6) | 1181 (4.8) | 5.2 |
| South Asian | 768 (14.2) | 4200 (10.3) | 4968 (10.7) | 17.6 | 503 (15.2) | 2354 (11.1) | 2857 (11.7) | 19.1 |
| Middle Eastern | 130 (2.4) | 885 (2.2) | 1015 (2.2) | 4.9 | 82 (2.5) | 459 (2.2) | 541 (2.2) | 4.6 |
| New Zealand Maori | 64 (1.2) | 372 (0.9) | 436 (0.9) | 0.6 | 30 (0.9) | 144 (0.7) | 174 (0.7) | 0.5 |
| Pacific Islander | 70 (1.3) | 365 (0.9) | 435 (0.9) | 0.9 | 29 (0.9) | 128 (0.6) | 157 (0.6) | 1.0 |
| Other | 380 (7.0) | 2515 (6.2) | 2895 (6.3) | 26.8 | 159 (4.8) | 967 (4.5) | 1126 (4.6) | 26.3 |
| *Unknown/Missing* [f] | 1084 (19.2) | 8417 (19.9) | 9501 (19.8) | 1.8 | *612 (17.9)* | *4231 (19.4)* | *4843 (19.1)* | *1.6* |
| Aboriginal and/or Torres Strait Islander [g] | | | | CCOPMM [d] | | | | ABS [e] |
| Yes | 74 (1.9) | 512 (1.4) | 586 (1.4) | 1.6 | 25 (1.1) | 184 (1.0) | 209 (1.0) | 0.7 |
| No | 3862 (98.1) | 36 848 (98.6) | 40 710 (98.6) | - | 2344 (98.9) | 18 642 (99.0) | 20 986 (99.0) | 99.3 |
| *Unknown/Missing* [f] | *1696 (30.1)* | *4924 (11.6)* | *6620 (13.8)* | - | *1057 (30.8)* | *3031 (13.9)* | *4088 (16.2)* | *0.2* |
| Language Spoken at Home [h] | | | | ABS [e] | | | | ABS [e] |
| English only | 3524 (69.4) | 28 127 (75.0) | 31 651 (74.3) | 59.1 | 2167 (71.3) | 14 448 (76.4) | 16 615 (75.7) | 60.6 |
| Mandarin | 123 (2.0) | 1 269 (2.8) | 1 392 (2.7) | 5.1 | 67 (1.8) | 592 (2.6) | 659 (2.5) | 4.5 |
| Punjabi | 273 (4.4) | 1082 (2.4) | 1355 (2.6) | 4.1 | 164 (4.4) | 503 (2.2) | 667 (2.5) | 4.6 |
| Hindi | 186 (3.0) | 1137 (2.5) | 1323 (2.6) | 2.4 | 126 (3.4) | 677 (3.0) | 803 (3.0) | 2.7 |
| Vietnamese | 159 (2.5) | 518 (1.1) | 677 (1.3) | 2.5 | 78 (2.1) | 211 (0.9) | 289 (1.1) | 1.7 |
| Cantonese | 61 (1.0) | 488 (1.1) | 549 (1.1) | 1.1 | 34 (0.9) | 225 (1.0) | 259 (1.0) | 1.2 |
| Sinhalese | 74 (1.2) | 428 (1.0) | 502 (1.0) | 1.1 | 49 (1.3) | 237 (1.0) | 286 (1.1) | 1.3 |
| Arabic | 60 (1.0) | 402 (0.9) | 462 (0.9) | 2.3 | 25 (0.7) | 147 (0.6) | 172 (0.6) | 2.0 |
| Spanish | 45 (0.7) | 365 (0.8) | 410 (0.8) | 0.7 | 25 (0.7) | 207 (0.9) | 232 (0.9) | 0.7 |
| Urdu | 61 (1.0) | 298 (0.7) | 359 (0.7) | 1.4 | 36 (1.0) | 137 (0.6) | 173 (0.6) | 1.5 |
| *Unknown/Missing* [f] | *546 (9.7)* | 4728 (11.2) | 5274 (11.0) | 1.0 | 387 (11.3) | *2931 (13.4)* | *3318 (13.1)* | 0.7 |

[a] Parent who gave birth to the index child including non-binary and transgender parents.
[b] Non-birthing parents who identify as male.
[c] Countries listed are only the ten most frequently reported non-English speaking countries for comparison with population data.
[d] Consultative Council on Obstetric and Paediatric Mortality and Morbidity (CCOPMM) aggregate data for births in 2021.
[e] Australian Bureau of Statistics (ABS) 2021 Australian Census, up to 42-year-old female parents and 45- year-old male parents.
[f] Percentage of total with unknown/missing data; remaining percentages within each category were calculated from only those with available data.
[g] Respondents could select multiple ethnicities including Aboriginal and/or Torres Strait Islander. GenV respondents were asked if they identified as Aboriginal and/or Torres Strait Islander in a second stand-alone item, leading to slightly different frequencies. Other ethnicities included Australian, New Zealand Peoples, South/Central/North American, Caribbean Islander, and unspecified; ABS comparison obtained from Ancestry question, noting a large proportion of “Other” had selected “Australian”.
[h] Languages listed are only the ten most frequently reported by birth mothers/parents.
Dash (-) = Data are not available; NA = Not applicable. Cells ≤ 10 are confidentialised.
